# Assessing the Impact of Dental Malocclusion on the Body’s Postural Balance: Correlation between Angle Class, Pelvic Balance, and Center of Foot Pressure

**DOI:** 10.1101/2023.12.21.23300397

**Authors:** Imad Akensous, Samia Karkouri, Chaimae Iziki, Wiam Rerhrhaye, Anissa Abdelkoui

**Author notes:** These authors contributed equally to this work.

## Abstract

**Background:** The purpose of this study was to determine the association between pelvic and center foot pressure (CoP) imbalances and angle class II or III malocclusions.

**Methods:** Our study is a prospective, descriptive, and analytic study conducted on two groups: a test group of 53 patients who present malocclusion class II or III compared with 53 controls matched for age and gender. The evaluation of the center of foot pressure (CoP) and the confidence ellipse area (mm2) were performed by stabilometric platefrom using bipodal test in two occlusal conditions, in maximum intercuspation (MI) and with a cotton roll (CR), with and without visual cue. A pelvic level device was used to perform the pelvic balance examination. Statistical analysis used the chi-square test, the McNemar test, and the Pearson test.

**Results:** In the case group, the bipodal test was poor (outside reference values) in MI open eyes, MI closed eyes, CR open eyes, and CR closed eyes in 47.2%, 62.3%, 58.5%, and 64.2%, respectively, vs 54.7%, 43.4%, 34.0%, and 67.9%, respectively, of controls (p <0.05). None subjects in the control group were diagnosed with pelvic imbalance, against five patients (9.4%) in the case group (p <0.05). Pelvic imbalance was noted in two patients in class II and only one patient in class III. For Class II, the bipodal test results were poor in MI open eyes, MI closed eyes, CR open eyes, and CR closed eyes conditions with percentages of 54.2%, 66.7%, 70.8%, and 58.3%, respectively (p <0.05). In Class III, results were generally good in MI open eyes (80%), but mostly poor in MIP and CR closed eyes (90%).

**Conclusion:** There was a significant difference in pelvic imbalance between the case group and the control group. Angle Classes II and III had no significant correlation with pelvic tilt. The stabilometric examinations have shown that Angle class II influences the CoP displacement and the confidence ellipse area in MI open eyes conditions. This finding supports the hypothesis that dental malocclusion can have an effect on the postural system.

## Introduction

An ocular motor problem, a hearing loss, or an unstable podal support systematically influences postural balance; on the other hand, a problem with the manducatory apparatus, such as a dental malocclusion, could have an impact on postural stability. In fact, if the podal sensor, the visual system, and the vestibular system are classically considered as postural sensors that participate in one way or another in postural control, the manducatory apparatus is not yet classified as a postural sensor.

In recent years, many studies have focused on the potential correlations between the stomatognathic system and body posture. Several biomechanical and neurophysiological hypotheses have attempted to explain this correlation, such as muscle chain, trigeminal nerve activation or inhibition, sterno-cleido-mastoidian muscle contraction, and facial chain theory [1] [2] [3]. A study by Amaricai E (2020) showed that there are some differences between different mandibular positions in subjects with physiological occlusion; a maximum mouth opening (no dental contact) deteriorates the static balance, whereas the postural stability improved in maximum intercuspidation compared to the mandibular postural position [4]. Sakaguchi in 2007 studied the effect of different mandibular positions on postural stability and found that the body is more stable when the subject is in centered occlusion than when the mandible is in resting or eccentric position (smaller center of pressure). This change in mandibular posture affects the cervical and facial muscles via the trigeminal nerve [5]. Using a stabilometric table, Tardieu et al. concluded that posture is only affected by occlusal dysfunction in the dynamic state (when the patient starts to move or walk) and not in the static state [6]. Several authors will reach the same conclusion as Sakaguchi and Tardieu. Like them, Wakano in 2011, and Tingey in 2003, studied the effects of voluntary lateral deviation of the mandible on postural balance and deduced that changes in the stomatognathic system significantly affect postural balance [7] [8]. Hanke, in his systematic review of 355 articles, concluded that there is a clear clinical link between dental occlusion and posture, but a causal link is still difficult to prove [9]. Amat in 2009 and Gasq in 2010 did comparable work and reached a similar conclusion [10] [11].

In contrast to previous findings, Ferrario reached completely different results and concluded that the foot pressure center is not affceted by any of the mandibular positions [12]. In three of his studies, Perinetti attempted to determine the relationship between occlusion and posture using posturography [13] [14] [15]. There is a significant difference in displacement of the pressure center between closed eyes and open eyes conditions, but no significant difference between the two positions of the mandible (rest position and maximum intercuspid position) has been detected. The author concludes that posturography is currently unable to detect this correlation.

In 2006, Michelotti demonstrated that only changing the position of the temporomandibular joint affects postural balance, but not dental occlusion [16]. According to März (2017) it is not possible to show a relationship between occlusion and posture, since the body compensates immediately at the neuromuscular level after the change of mandibular position to adapt and find a new balance [17]. A recent systematic review of major databases showed that 66.7% of articles showed an association between dental occlusion and body posture, and 33.3% found no relationship [18]. However, the majority of articles published are based on protocols study of low methodological value that are generally done on small groups of subjects without a control group and carried out on healthy subjects not suffering from any dental malocclusion [19].

The impact of occlusal problems on postural control remains controversial, as most of the information available to date is inconclusive and recent reviews have reported contrasting results. In this heated debate, among the scientific communities, it is difficult to reach to a unanimous conclusion.

The purpose of our study was to demonstrate, through a case-control study the existence of a statistically significant association between Angle class malocclusions and pelvic imbalance, Angle class malocclusions and center of foot pressure displacement depending on two dental and visual conditions.

To our knowledge, no clinical case-control study has been conducted to investigate the probable correlation between dental malocclusion and postural imbalance on a national and continental scale.

## Material and methods

### Subjects

This prospective descriptive-analytical case-control study was conducted among 106 individuals divided into two equal groups of 53 patients. The sample of cases with malocclusion was recruited from the dental consultation and treatment center at the department of Dentofacial Orthopedic, while the 53 healthy subjects were recruited from students of Faculty of Dentistry, their relatives, and acquaintances.

We calculated the sample size formula using “riskcalc-simple size calculator” software [20]. A sample of subjects with malocclusion will be compared for postural balance with a sample of controls. Assuming an equal number of cases and controls (i.e., k = 1). For achieving an 80% power (i.e., 1 − β = 0.8) at the 5% level of significance (i.e., α = 0.05) and for p0 = 0.66, the sample size is 46 cases and 46 controls or 53 cases and 53 controls by incorporating the continuity correction [21] [22] [32].

All subjects who participated in this study underwent their medical history examined by means of a questionnaire, followed by an intraoral and extraoral examination to detect any possible oral pathology except dental malocclusion. This study included patients aged from 14 to 55 years, of both sexes. The selecting criteria for the test group were: good general health according to anamnesis and through clinical examination; complete dentition and Angle class II or III. On the other hand, the control group had any dental malocclusion traits but with a bilateral Angle class I of molars and canines. The exclusion criteria were: an age below 14 years and above 55 years; partially or completely toothless; temporomandibular dysfunction (TMD) or any other disorder affecting the mandibular apparatus; vestibular or sensory disease; ongoing orthodontic treatment; prior or ongoing orthopedic treatment; acute or chronic orofacial or vertebral inflammatory diseases and subjects who performed an orthognathic surgery.

All patients were referred to the Dentofacial Orthopedic service, then to the Physical Medicine and Rehabilitation department during the period ranging from 10 October 2022 to 21 May 2023. The experimental protocol complies with the Declaration of Helsinki on ethical principles for research involving human subjects and it was approved by the local Ethics Committee (59/2022). A written informed consent form was obtained from each patient or their legal guardian before the tests.

### Methods

#### Angle class assessment

The examination of the right and left canine and molar dental angle class was performed by an experienced orthodontist directly on the oral cavity; then recorded on a database. Examiner reliability was confirmed by repeated intra-examiner examinations for 50 subjects. The intra-examiner ICC was about 0, 8.

Angle Class I, II or III are evaluated based on the positioning of the first molar and canine between the two arches. If the maxillary canine is opposite the embrasure between the mandibular canine and the lower 1st premolar, and the mesiobuccal cusp of the maxillary 1st molar is aligned with the center (mesiobuccal groove) of the mandibular 1st molar, the patient is classified as Angle Class I [23].

Class II malocclusion is characterized by a distal position of the mandibular arch from its normal position, the mandibular first molar as well as the rest of the dentition is distanced from its normal position in relation to the maxillary first molar [24]. Class III malocclusion is characterized by distocclusion of the maxillary first molar from the mandibular first molar and of the maxillary canine from the mandibular canine [25].

#### Postural balance assessment

##### Assessment of pelvic balance

This examination is performed by a pelvic level device (Advanced Casting Technology ™, Boise, USA), an easy tool to use by the clinician for a quick and accurate assessment of the symmetry of the iliac crest. This device is composed of three expandable arms, in each one a level bubble [26].

To perform the test, the patient is asked to stand up, then the expandable wings of the tool are pressed on the pelvic bone crest, and by a simple observation of the bubbles, the pelvic balance is evaluated. When the three bubbles are in the center of the three branches, the pelvis is balanced; if not, it is deviated (S1 Fig).

#### Posturographic assessment

Postural stability is assessed by means of a sophisticated balance system (TecnoBody S.r.l., Prokin 252, Dalmine BG, Italy).This device consists of a stabilometric platform, a personal computer, and a 20-inch touch screen display. For correct feet position, the platform is marked with reference lines and is composed of three highly sensitive load cells and a trunk sensor to accurately obtain information about the patient’s balance during various tests. The stabilometric data were sampled at 20 Hz [27] [28].

The system contains several tests; we have chosen the bipodal test, which is conducted in a static situation and provides information about the displacement of the center of pressure (CoP) and about the confidence ellipse area (mm2) from reference values in two visual conditions (open eyes and closed eyes).

This test is recorded under two occlusal conditions: a first recording in MI and a second recording with a cotton roll (CR) of 10 mm thickness placed between the two central incisors of the two dental arches in order to eliminate the posterior periodontal receptors information and keep only that of the central incisors.

The practitioner places the subject precisely on the platform according to the following conditions: barefoot, a vertical position with the arms along the body without voluntary movements, soles of feet must be attached to the platform, and the examination room must remain calm throughout the test. If any of these conditions are not me, the test is canceled and then repeated. The entire examination is conducted by the same experienced practitioner.

At the first recording, the subject is asked to keep the teeth in maximum intercuspal position (MIP), to maintain a static standing position and to initially focus (eyes opened) on a “plus sign” displayed on the device’s screen before closing his eyes during the second part of the test (S2 Fig).

The recording takes 60 seconds (30 seconds with open eyes and 30 seconds with closed eyes). After a rest period of about 3 to 4 minutes (to avoid the effect of fatigue), a cotton roll is placed between the central incisors of the two dental arches, the subject is placed back on the platform exactly in the same way as the first recording, and then the data is recorded again for the same duration. Finally, the cotton roll is removed, and the subject leaves the platform. The measurements recorded in MIP and with a CR were taken consecutively in the same order.

The test results vary between three intervals represented by three colors: Green reflects that the CoP is included in physiological range and the confidence ellipse area is inside high quality reference values, yellow indicates a medium results but always in physiological range, while the orange color means that the CoP is not physiological and the confidence ellipse area is outside reference values.

#### Statistical analysis

In our study, the Angle class was analyzed with age, sex, and two postural parameters, which are pelvic imbalance and the bipodal test in two visual conditions (open eyes and closed eyes) and two occlusal conditions (MIP and CR).

Descriptive statistics included the mean and standard deviation for quantitative variables with a normal distribution, the median and interquartile for Gaussian distribution, the frequency and percentage for categorical variables. Comparisons between two groups, for categorical variables, were performed by the *χ^2^* test if the theoretical number is ≥ 5 or by the *Fisher test* if the observed number is <5, with 95% confidence interval (CI) calculated where appropriate. A comparison of a qualitative variable within the same group was performed by *McNemar test*. Correlation analysis was performed by *Pearson Chi-2 test* to assess the independent association of Angle class with the two posture variables. The p-value was bilateral and considered statistically significant if it was less than 0. 05. All analyses were performed using SPSS, version 24.0 (SPSS Inc., Chicago, IL).

## Results

This study was conducted on a sample of one hundred six patients: 57 males (53.77%) and 49 females (46.23%) aged between 14 and 55 years, median age 25 [21–36] years, divided into two groups of equal size. Table 1 represents the description of different variables in the case group.

**Table 1.**
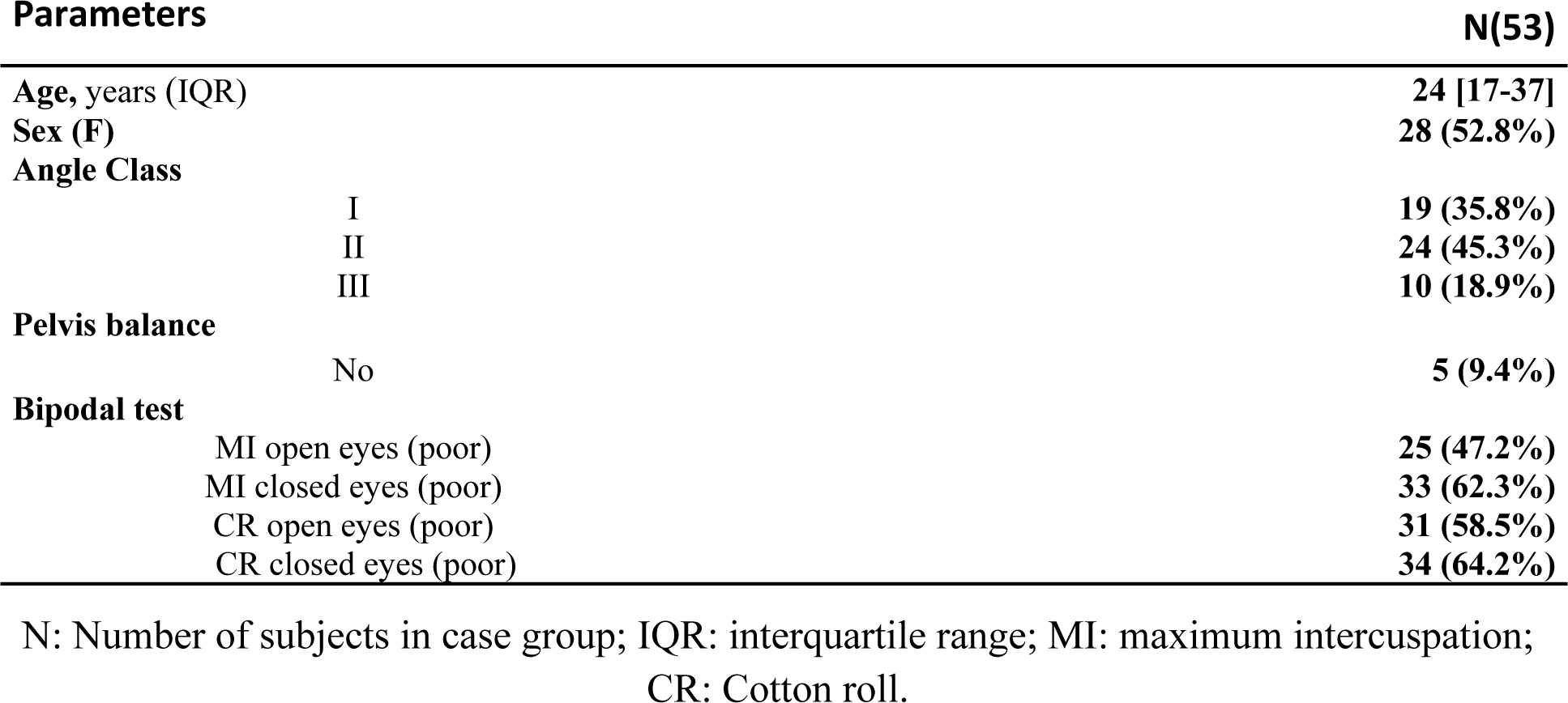
description of variables in the cases group (N = 53)

This group was composed of 53 patients (28 females and 25 males, mean age 24 [17–37] years). Analysis of the dental occlusion noted was as follows: 35.8% Angle class I, 45.3% class II and 18.9% class III. Pelvic imbalance was observed in 9.4% of subjects. The bipodal test was poor in MI open eyes, MI closed eyes, CR open eyes, and CR closed eyes for 47.2%, 62.3%, 58.5%, and 64.2%, respectively.

Description of different variables in the control group is shown in Table 2, which also consisted of 53 patients, (25 females and 28 males, mean age 26 [22–38] years). All subjects in this group belonged to Angle class I. No pelvic imbalance was noted. The bipodal test results were poor in the MI open eyes, MI closed eyes, CR open eyes, and CR closed eyes conditions for 54.7%, 43.4%, 34.0%, and 67.9% of subjects, respectively.

**Table 2.**
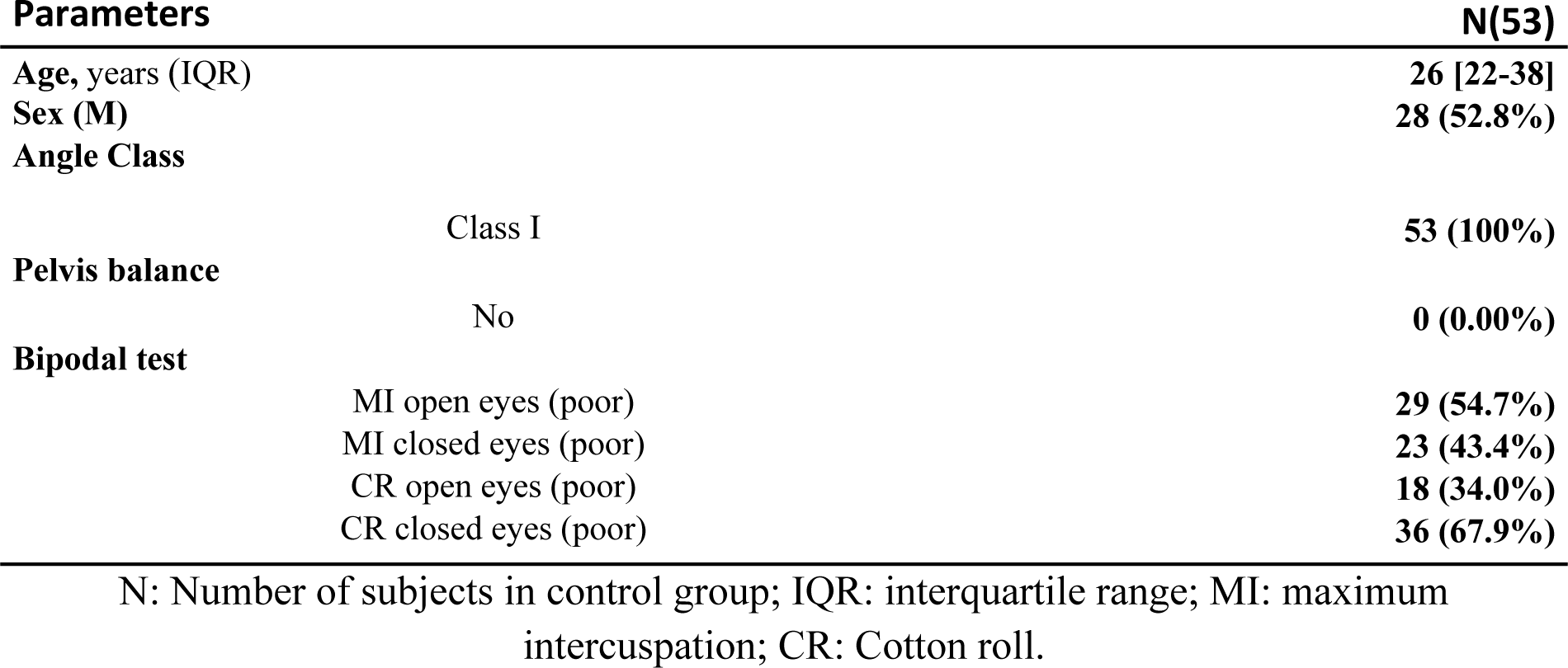
description of variables in the control group (N = 53)

The comparison of pelvic imbalance between the case and control groups is shown in Table 3.

**Table 3.** Comparison of pelvic imbalance between the case and control group.

| | | Case group | | Control group | | Total | | $\chi^2$ test |
| --- | --- | --- | --- | --- | --- | --- | --- | --- |
|  |  | Effective | Percentage % | Effective | Percentage % | Effective | Percentage % |  |
| Pelvic balance | Yes | 48 | 90.9 | 53 | 100 | 101 | 95.4 | **0.022\*** |
|  | No | 5 | 9.4 | 0 | 0.00 | 5 | 4.6 |  |

$\chi^2$ test: Chi-squared test ;\*: p-value significant if < 0.05.

No subjects in the control group were diagnosed with pelvic imbalance, against five patients (9.4%) in the case group.

The results of the bipodal test (CoP assessment) in visual and occlusal conditions between the two groups are represented in Table 4.

**Table 4.** Comparison of the Bipodal test between the two occlusal and visual conditions.

|  |  |  | MIP |  | CR |  | Mc-Nemar Test |
| --- | --- | --- | --- | --- | --- | --- | --- |
|  |  |  | Open eyes | Closed eyes | Open eyes | Closed eyes |  |
| Bipodal Test | Case group | Ex | 7 (13.2%) | 15 (28.3%) | 6 (11.3%) | 16 (30.2%) | NS |
|  |  | G | 21 (29.6%) | 5 (9.4%) | 16 (30.2%) | 3 (5.6%) |  |
|  |  | P | 25 (47.2%) | 33 (62.3%) | 31 (58.5%) | 34 (64.2%) |  |
|  | Control group | Ex | 14 (26.4%) | 30 (56.6%) | 14 (26.4%) | 14 (26.4%) | **0.01\*<0.05** |
|  |  | G | 10 (18.9%) | 0(0.00%) | 21 (39.6%) | 3 (5.7%) |  |
|  |  | P | 29 (54.7%) | 23 (43.4%) | 18 (34.0%) | 36 (67.9%) |  |

Ex : Excellent ; G : Good ; P : Poor ; NS : difference not statistically significant ; MIP : maximum intercuspation position ; CR : Cotton roll ; \*: p-value significant if< 0.05

The bipodal test results in the case group in MIP open eyes were physiological for 28 patients, i.e., 52.8%, compared to 41.5% in CR open eyes. In MIP closed eyes, 20 patients (37.7%) had normal results compared to 19 patients (35.8%) in CR closed eyes. In the control group, 45.3% of subjects performed good or excellently on the bipodal test in MIP open eyes, compared to 66% in CR open eyes conditions. In MIP closed eyes, 56.6% had physiological results vs 32.1% in CR closed eyes.

In the bipodal test, 26.4% of the control subjects had excellent results in MIP open eyes, against only 13.2% of subjects in the case group, whereas in MIP closed eyes, 28.3% of the cases had an excellent result against 56.6% of the controls. In CR open eyes conditions, 58.5% of patients with dental malocclusion (class II or III) had poor results vs 34% of the control group (class I), while with closed eyes, the results were relatively similar between the two groups.

Table 5 shows the correlation between Angle Class II and III, the bipodal test in two occlusal conditions, and the pelvic imbalance.

**Table 5:** Correlation between Angle class, bipodal test, and pelvic imbalance.

|  |  | Bipodal test |  |  |  | Pelvic balance |  |
| --- | --- | --- | --- | --- | --- | --- | --- |
|  |  | ICP |  | CR |  | No | Yes |
|  |  | Open eyes | Closed eyes | Open eyes | Closed eyes |  |  |
| Class II | Ex | 8.30% | 20.80% | 4.20% | 33.30% | 2 (8.3%) | 22(91.7%) |
|  | G | 37.50% | 12.50% | 25.00% | 8.30% |  |  |
|  | P | 54.20% | 66.70% | 70.80% | 58.30% |  |  |
| Class III | Ex | 0.00% | 10.00% | 0.00% | 10.00% | 1 (10%) | 9(90%) |
|  | G | 80.00% | 20.00% | 50.00% | 10.00% |  |  |
|  | P | 20.00% | 70.00% | 50.00% | 80.00% |  |  |
| Pearson correlation test |  | 0.018*<0.05<br>(Classe II) | 0.10 | 0.068 | 0.41 | 0.11 |  |

Ex : Excellent ; G : Good ; P : Poor ; ICP : intercuspal position ; CR : Cotton roll ; \*: p-value significant if< 0.05.

Pelvic imbalance was diagnosed in two patients with class II and only one patient with class III. For Class II, the bipodal test results were poor in MIP open eyes, MIP closed eyes, CR open eyes, and CR closed eyes conditions with percentages of 54.2%, 66.7%, 70.8%, and 58.3%, respectively. Class III had generally a good result in MIP open eyes (80%), but mostly poor in MIP and CR closed eyes (90%).

## Discussion

The aim of this study was to investigate the supposed correlation between dental malocclusion, especially class II and III malocclusion, with pelvic imbalance and CoP displacement. A clinical examination of the pelvic girdle, as well as objective posturographic tests under different occlusal and visual conditions was performed, for the case group (dental malocclusion) and the control group (normocclusion).

In this study, we reported a significant difference between the case group and the control group regarding to pelvic imbalance. It was significantly higher in subjects with malocclusion compared to controls. These findings suggest that Angle Class II or III may be related with pelvic imbalance. A correlation analysis by pearson’s chi-2 test was performed to determine the association between Angle class malocclusions and pelvic tilt. This analysis showed that there was no significant correlation between class II or III and pelvic imbalance. A previous study conducted by Lippold in 2006, consisted of 53 healthy subjects (32 women, 21 men; mean age 24.6 years), six angular measurements of the skeleton by lateral head cephalography and an examination of the sagittal profile of the back by rasterography were performed, in order to analyze the probable relationships between spinal posture (thoracic, lordotic, and pelvic tilt) and the craniofacial morphology. They reported a statistically significant correlation between craniofacial parameters and a low back profile. Normal back parameters were higher in patients with a distal (Class II) and vertical mandible (upper thoracic, lumbar lordosis, pelvic angle), and vice versa for patients with a mesial and horizontal mandible (Class III) [29]. In 2007, the same author demonstrated a correlation between the facial axis and facial depth of subjects with Class II and III malocclusion with pelvic torsion by cephalometric and rastereographic analysis. Statistically significant differences in pelvic torsion were reported in relation to facial axis and facial depth. Patients with a vertical facial pattern and a distally positioned mandible have a slight pelvic torsion where the left iliac spine faces backward relative to the right iliac spine. Patients with a horizontal facial pattern and a mesially positioned mandible have a slight pelvic torsion where the right iliac spine faces backward relative to the left iliac spine. Therefore, the author concluded that an orthopedic examination can be considered for patients undergoing orthodontic treatment [30].

In the present study, we also performed a posturographic examination (the bipodal test) in two occlusal conditions, in intercuspal position (ICP) and with cotton roll (CR), in order to evaluate the oscillation of the CoP and the confidence ellipse area (mm2) with and without the visual cue. In the case group, there was no significant difference between the two occlusal conditions, suggesting that ICP or CR conditions did not affect either the CoP oscillation or the confidence area.

In contrast, there was a significant difference between ICP and CR in the control group. More subjects exhibited physiological results in the CR open eyes conditions compared to the ICP open eyes conditions. These findings suggest that CR improved the bipodal test performance of control subjects. However, the results were reversed in CR with closed eyes (visual sensor deactivated), showing a highly significant difference towards a CoP and a confidence area outside reference values. This suggests that the ocular sensor may have compensated for the effect of CR when the eyes are open. These results can be confirmed more in the ICP with closed eyes, where the significance is very high for a physiological oscillation of the CoP and the confidence area. On the basis of these results, we can say that the oscillation interval of the CoP and the confidence ellipse area improved in ICP closed eyes conditions (the best results were obtained in ICP closed eyes), which suggests that the physiological dental occlusion of controls compensated for the absence of the visual sensor, which defends the hypothesis that the dental occlusion can be considered as a main postural sensor. März et al., in their pilot study published in 2017, asked the question: “Can different occlusal positions instantaneously impact spine and body posture?” To answer this question, they selected 44 healthy volunteers and evaluated ten postural variables with four mandibular positions (right eccentric, physiological rest, cotton rolls on both sides, and 1 mm occlusal elevation) using rasterography. Significant differences were found for cervical spine, lumbar spine, and kyphotic angle with these different mandibular positions. However, they could not conclusively associate dental occlusion conditions to instantaneous body posture changes, since the postural variations could also come from neuromuscular compensation [17].

In this research, a significant difference was reported between the two groups, in the bipodal test, suggesting that there is a potential effect of dental malocclusion on the CoP and on the confidence area. A correlation analysis between the bipodal test, Class II and III dental malocclusion showed that Angle Class II significantly influences CoP and the confidence ellipse area in ICP with open eyes conditions. According to Baldini et al, the force platform is not able to detect clearly the relationship between dental occlusion and body posture. It has been shown that vision influences body posture during stabilometric platform testing and not mandibular positions [31]. Sakaguchi’s study reported a significant difference between the total length of the CoP path with different mandibular positions. The total CoP path length in centric occlusion was shorter than in the resting position. The CoP area in right eccentric mandibular position was larger than in centric occlusion. Based on these results, it was concluded that mandibular positions affect body posture [5].

The lack of comparison of the exclusive effects of the eye sensor on posture in stabilometric plateform evaluation between the case and the control group is a limitation of our study.

Overall, it would be interesting to conduct an additional study that will include other types of dental malocclusion with new postural parameters and to use posturographic tests in the static as well as dynamic state to reveal other possible effects of dental malocclusion on the postural system.

## Conclusion

In our study, we demonstrated through clinical examinations that there is a significant difference in pelvic imbalance between the case and the control group, but no significant correlation was noted between Angle Class II or III and pelvic tilt. Through the stabilometric examination (bipodal test), we demonstrated that Angle class II influences the CoP and the confidence ellipse in the ICP open eyes conditions. The same test proved that the normocclusion of control subjects improved or perfectly compensated the postural balance in case of absence of the ocular sensor (eyes closed). All of this, supports the hypothesis that dental malocclusion can have an effect on the postural system, as well as the postulate considering dental occlusion as a postural sensor.

## Data Availability

The data underlying this research will be fully accessible on Figshare upon article acceptance, with the assigned DOI being DOI : 10.6084/m9.figshare.24757050. This DOI will become active after the article has been accepted for publication.

https://figshare.com/account/articles/24757050

## Supporting information

**S1 Fig. Examination of the pelvis balance (iliac crest) using a “pelvic level”**

**S2 Fig. Photo of a patient during posturographic examination (bipodal test)**

## Notes

### Competing Interest Statement

The authors have declared no competing interest.

### Funding Statement

The author(s) received no specific funding for this work.

### Author Declarations

The research study conducted at the Faculty of Medicine and Pharmacy of Rabat received ethical approval from the institution's review board. The approval was granted with the reference number 59/22. To ensure ethical standards, all participating patients willingly and voluntarily signed a written and informed consent form prior to their involvement in the study. This process aligns with the guidelines and ethical considerations set forth by the Faculty of Medicine and Pharmacy of Rabat's ethics committee, ensuring the protection of participants' rights and well-being throughout the research endeavor

